# Deep brain stimulation improves symptoms of spasmodic dysphonia through targeting of thalamic sensorimotor connectivity

**DOI:** 10.1101/2022.11.26.22282419

**Authors:** Michael G Hart, Nancy Polyhronopoulos, Mandeep K Sandhu, Christopher R Honey

## Abstract

**Background:** Spasmodic dysphonia is a dystonia of the vocal chords producing difficulty with speech. Current hypotheses are that this is a condition of dysregulated thalamic sensory-motor integration. A recent randomised controlled trial of thalamic deep brain stimulation (DBS) demonstrated its safety and efficacy. Our objective was to determine if the outcome could be predicted by stimulation of thalamic sensorimotor regions and adjacent white matter connectivity as assessed by diffusion tractography.

**Methods:** A cohort of six participants undergoing thalamic deep brain stimulation for adductor spasmodic dysphonia was studied. Electrodes were localised with the Lead-DBS toolbox. Group-based analyses were performed with atlases, co-ordinates, and using voxel-based symptom mapping. Diffusion tensor imaging (3 Tesla, 64 directions, 2mm isotropic) was used to perform individual probabilistic tractography (cerebellothalamic tract and pallidothalamic tract) and segmentation of the thalamus. Monopolar review was performed at 0.5V and binarised as effective or ineffective.

**Results:** Effective contacts stimulated more of thalamic regions connected to sensorimotor cortex than ineffective contacts (p<0.05, FDR corrected). This effect was consistent across analytical and statistical techniques. Group-level and tractography analyses did not identify a specific ‘sweet spot’ suggesting the benefit of DBS is derived from modulating individual thalamic sensorimotor connectivity. Stimulations at one year involved predicted thalamic sensorimotor regions with additional cerebellothalamic tract involvement.

**Conclusions:** Symptoms of spasmodic dysphonia are effectively treated by DBS. Stimulation of thalamic sensorimotor areas was associated with symptomatic improvement. These data are consistent with DBS acting upon pathophysiologically dysregulated thalamic sensorimotor integration in spasmodic dysphonia.

**What is known on this topic:** Spasmodic dysphonia is a dystonia affecting speech with few treatment options other than speech therapy and botulinum toxin.

**What this study adds:** Deep brain stimulation is demonstrated to be an effective therapy by targeting sensorimotor areas of the thalamus.

**How might this study affect research, practice or policy:** These data add to the evidence that spasmodic dysphonia is due to dysregulated thalamic sensorimotor integration, and also suggest novel targets for steering stimulation towards to maximise benefit.

## INTRODUCTION

Spasmodic dysphonia, also known as laryngeal dystonia, is a task-specific focal dystonia of the laryngeal muscles that impairs speech. It is rare with an incidence of 1-4 per 100,000 per year and most often occurs in females aged 30 to 50 years. Subtypes include the more common adductor form (characterised by short staccato speech), the less common abductor form (characterised by whispering breathy speech), and finally the least common mixed adductor and abductor variant. Current hypotheses regarding its pathophysiology include disordered sensorimotor integration(1) and cortico-thalamic connectivity as well as dopamine receptor imbalance.(2)

Treatment options traditionally include speech therapy and botulinum toxin injections. A rare association of spasmodic dysphonia with tremor led serendipitously to the observation that thalamic DBS used to treat tremor also improves symptoms of spasmodic dysphonia.(3) A recent blinded, randomised controlled trial of thalamic deep brain stimulation for adductor spasmodic dysphonia demonstrated a sustained improvement at 12 months.(4) Questions are now focused on DBS and its mechanism of action, optimal targeting, patient selection, and predicting individual outcomes.

Our hypothesis was that DBS targeted the integration of sensorimotor connectivity within the thalamus. To test this, we used patient data from a recent phase 1 RCT of thalamic DBS for spasmodic dysphonia. Additionally, we performed tractography with diffusion MRI data to identify individual connectivity of the thalamus and surrounding white matter. Our aims were to understand how DBS interacted with spasmodic dysphonia pathophysiology within the thalamus and define individual biomarkers of response that could be used for targeting.

## MATERIALS AND METHODS

### Ethical approval

This single centre, prospective cohort study received ethical permission from The University of British Columbia Clinic Research Ethics Board (H15-02535). The trial protocol was prospectively registered on clinicaltrials.gov (NCT02558634). Written informed consent was obtained in line with the Declaration of the Principles of Helsinki.

### Recruitment

A cohort of people with refractory symptoms of adductor spasmodic dysphonia attending specialist clinic follow-up were invited to participate. Diagnosis of isolated adductor spasmodic dysphonia (i.e. without associated tremor) was by consensus among two laryngologists and a specialist speech language therapist.

### Neurosurgery

Deep brain stimulation was performed in the awake patient using an MRI-guided CT-verified procedure utilising the Cosman-Roberts-Wells (CRW®) frame (Integra® LifeSciences, Cincinnati OH, USA). All patients had a single Medtronic 3389 electrode implanted into their left thalamic Vim (figure 1) and connected to an Activa SC implantable pulse generator (Medtronic Inc, Dublin, Eire). Targeting was performed indirectly on the Medtronic StealthStation S8 using AC-PC co-ordinates according to the following principles: lateral 10mm from border of the 3^rd^ ventricle, anterior 25% of the AC-PC distance posterior to the MCP, and vertical in the AC-PC plane. Intra-operative physiological confirmation of targeting was performed with macrostimulation (TC-16-2-250-D electrode and Cosman G4 generator; Boston Scientific Corporation, Marleborough, Mass, USA) at a frequency of 50Hz and pulse width of 1ms until paraesthesia was obtained at 0.8 to 1.5V without internal capsule effects. Post-operative CT images were acquired the following day.

**Figure 1:**
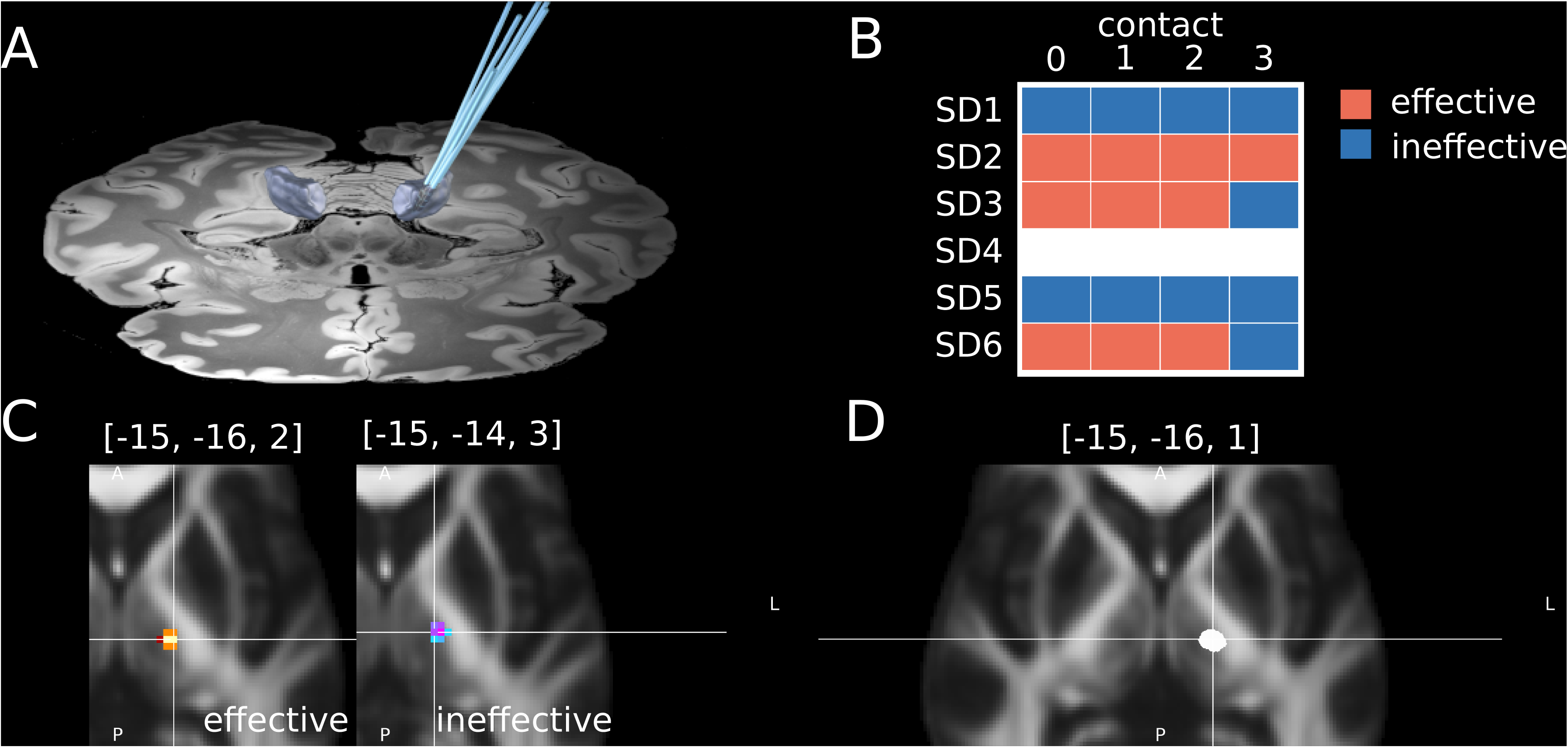
Study Methodology. A: Electrode (Medtronic 3389) reconstruction from Lead-DBS with the thalamic Vim nucleus from the DISTAL atlas highlighted in grey B: Monopolar review for 5 out of 6 participants highlighting effective and ineffective contacts on symptom relief at 0.5V C: Group overlay of effective (red, left) and ineffective (blue, right) contacts and their VATs at 0.5V together with centre-of-gravity co-ordinates. D: Final clinical stimulation VATs for all 6 participants. All VATs were computed using the finite element methods within Lead-DBS.

### MRI acquisition

All participants underwent MRI scanning at 3 Tesla using a Siemens Magnetom Prisma-fit scanner and 16-channel receive-only head coil (Siemens AG, Erlangen, Germany) with protocols based on those of the Human Connectome Project(5). Sequences included MPRAGE and diffusion tensor imaging (2mm isotropic, 64 directions).

### Electrode localisation

Registration with ANTs(6) and electrode reconstruction with PaCER(7) was performed using the Lead-DBS toolbox(8) as previously described.(9) Stimulations were simulated for each monopolar contact at 0.5V using the in-build algorithm for computing the volume of activated tissue (VAT) in Lead-DBS (figure 1). Atlases including the DISTAL(10), Human Motor Thalamus(11), and FGATIR hypointensity(12) were then used as overlays for VATs.

### Tractography analysis

Diffusion MRI data was processed (figure 2) using FSL tools(13) including BedPostX and ProbtrackX.(14) Tractography was performed for the cerebellothalamic tract (CTT) (seed: ipsilateral dentate(15), way masks: red nucleus and Vim (Distal) and pallidothalamic tract (PTT) (seed: ipsilateral pallidum (DISTAL), exclusion masks: retrolenticular and anterior limb of internal capsule(16) and striatum (DISTAL), targets: VIM and substantia nigra (DISTAL)). Thalamic segmentation was performed by defining a mask of the thalamus with FIRST(17) which was subsequently used as a seed for tractography. Delineation of individual thalamic segments was performed in three ways: using clusters based on individual cortical targets (FreeSurfer(18) defined primary motor (region 21), primary sensory (region 23), secondary motor (region 27), and contralateral dentate nucleus(15) set at a cluster threshold of 1000); hard-segmentation approach into areas of dominant connectivity(14) to individual cortical networks defined on the Yeo template(19); and a data-driven k-means segmentation (set at 4 clusters) approach to individual cortical grey matter (defined with ANTs) using k-means clustering.

**Figure 2:**
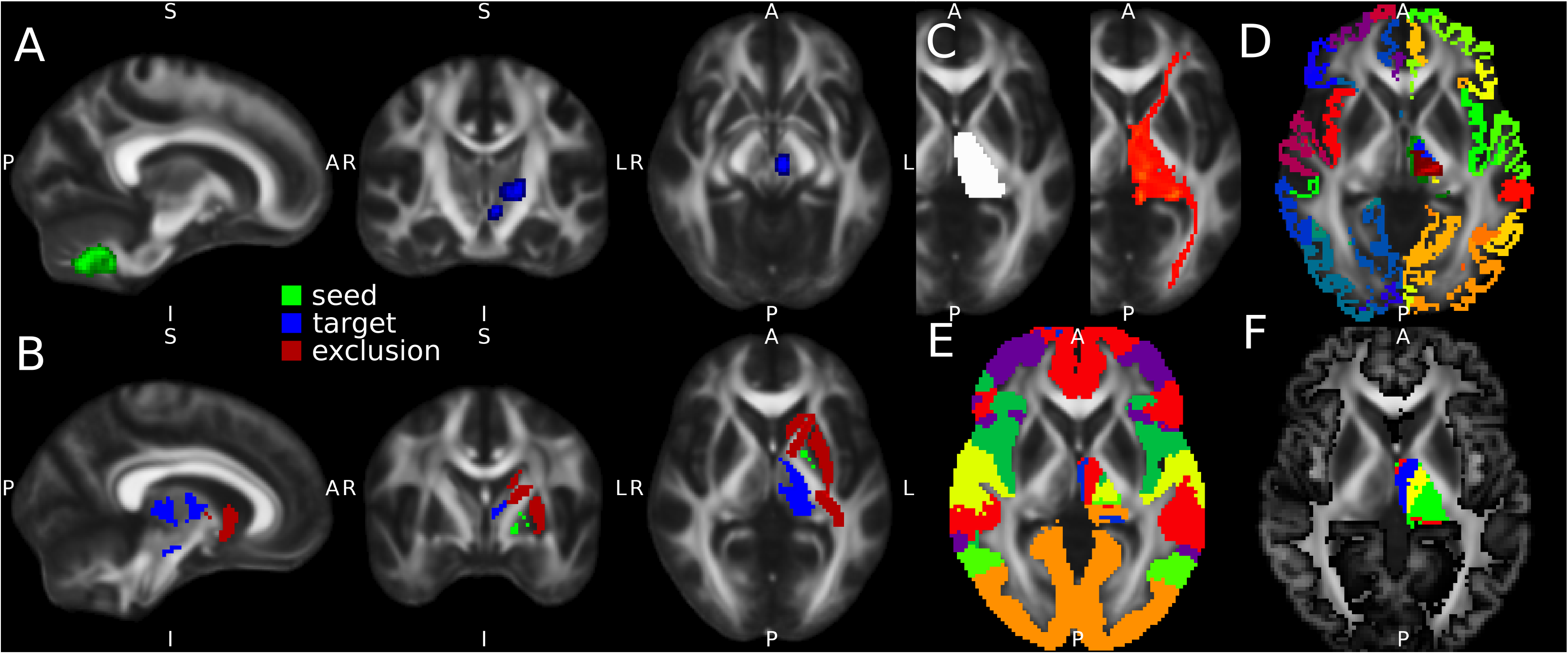
Neuroimaging Analyses. A: The cerebello-thalamic tract (CTT) was reconstructed from the dentate nucleus with waymasks through the contralateral red nucleus and thalamic Vim. B: The pallido-thalamic tract (PTT) was reconstructed from the pallidum to the thalamic Vim and substantia nigra with exclusion masks in the anterior and posterior limbs of the internal capsule as well as the putamen. C: A mask of the thalamus (left side of image) was used as a seed for probabilistic tractography (right side of image). D: clustering was performed based on connectivity to individual cortical targets (primary motor, primary sensory, secondary motor) derived from FreeSurfer and the contralateral dentate nucleus. E: a winner-takes-all approach of connectivity to cortical targets derived from the Yeo atlas. F: data-driven k-means segmentation to an individual cortical mask derived from ANTs.

### Study design

All assessments were performed by two specialists and single blinded to the participant. Firstly, a monopolar review of individual contacts was performed at 0.5V defining outcomes as either ‘effective ‘or ‘ineffective’ on SD symptoms (including clarity, fluency, pauses, and volume). This threshold was identified as the lowest voltage able to objectively demonstrate a consistent clinical effect (figure 1). Frequency (180Hz) and pulse width (60us) were kept constant throughout. Additionally, clinical outcomes at 12-month follow-up were recorded for voice-related quality of life (V-RQOL) and voice handicap index (VHI) together with the clinical stimulation parameters used at that time.

### Statistics

Statistical analyses were performed in MATLAB (The MathWorks Inc, Nantucket, Massachusetts). For the monopolar review, voxel-based analysis was performed using chi-square test of independence on effective versus ineffective contacts. Template and diffusion MRI biomarker analyses were performed with paired t-tests assessing overlap of VATs with selected biomarkers (standard space atlases, CTT, PTT, and all 3 thalamic segmentation methodologies). Clinical outcomes at 12 months were tested with diffusion MRI biomarkers using Pearson’s product-moment correlations. All analyses were FDR-corrected according to Benjamin and Hochberg principles with a significance level set at *p*<.05.

### Data Availability

Anonymised data will be made available upon reasonable request.

## RESULTS

### I. Demographics and clinical outcomes

A total of six participants were recruited whose clinical results of thalamic DBS are previously reported (table 1). A single participant’s data was excluded from monopolar review due to logistical issues. All participants demonstrated improvement in V-RQOL, the trial’s primary outcome measure, although this was not statistically significant.

**Table 1:** Patient Characteristics

| Participant |  | 1 | 2 | 3 | 4 | 5 | 6 |
| --- | --- | --- | --- | --- | --- | --- | --- |
| Stimulation | Contacts | -+00 | C+/-000 | C+/0-00 | C+/0-00 | + -00 | C+/0-00 |
|  | Frequency | 185 | 185 | 185 | 185 | 185 | 185 |
|  | Pulse width | 90 | 90 | 90 | 60 | 60 | 60 |
|  | Voltage | 1.5 | 2.1 | 3.5 | 2.0 | 1.3 | 2.6 |

Contact nomenclature is: 0123 distal-to-proximal, - = cathode, + = anode, 0 = no stimulation, C = case

### II. Group-level analyses do not identify ‘sweet spots’ of benefit

To assess for a group-level ‘sweet spot’ for stimulation we performed three analyses (figure 3). Firstly, we tested whether effective contacts stimulated more of putative targets in template atlases than ineffective contacts. The volumes stimulated for each of our template atlases – including the VIM (effective: M 3.82 SD 0.47, ineffective: M 4.16 SD 0.95, t(18)=-1.02, *p*=.32), VLdVLv (effective: M 1.52 SD 0.23, ineffective: M 1.55 SD 0.41, t(18)=-0.23, *p*=.082), and FGATIR hypointensity (effective: M 3.79 SD 3.52, ineffective: M 1.79 SD 3.01, t(18)=1.35, *p*=.019) – the results were equivocal. Next, we performed a voxel-wise stimulation-symptom mapping approach which did not identify any significant voxels of benefit (*χ*^2^, *p*>.2). Finally, we tested whether specific Cartesian (XYZ) co-ordinates were associated with effectiveness. In this latter analysis, while a more negative Y co-ordinate (i.e. more posterior) was associated with effective contacts (effective: M -15.97 SD 1.36, ineffective: M -14.23 SD 1.25, t(18)=2.97, *p*=.008), this was not significant when corrected for multiple comparisons.

**Figure 3:**
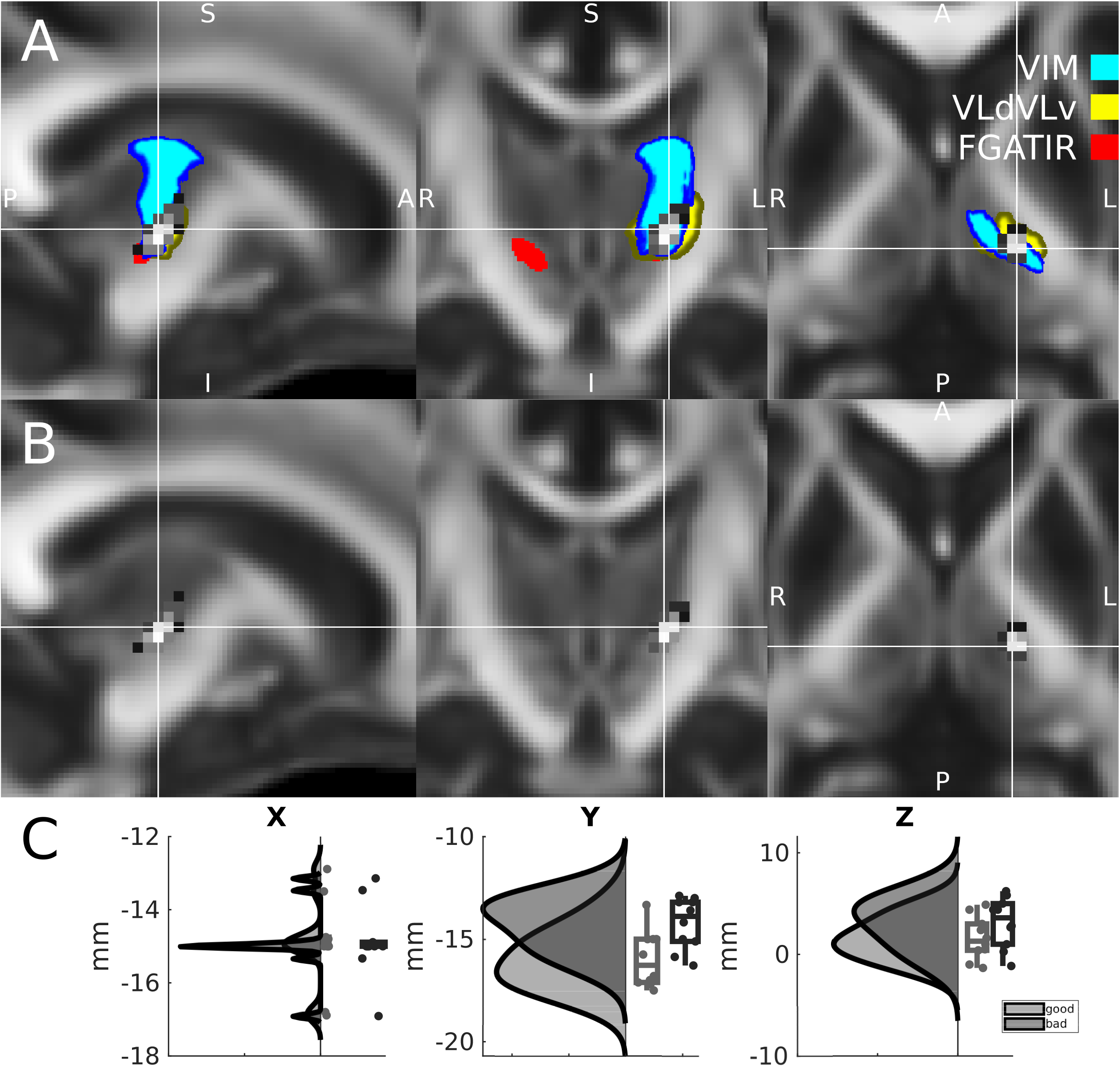
Group-based Analysis of Atlases, Individual Voxels, and Co-ordinates. A: An overlay of all monopolar VATs (grey) with atlas templates. B: voxel-based stimulation symptom mapping was performed for effective versus ineffective contacts. C: Raincloud plots (a composition of raw data, boxplots of mean and confidence intervals, and half-violin plots reconstructing the data probability density) of Cartesian XYZ co-ordinates. No significant effect was demonstrated after correction for multiple comparisons.

### III. Effective contacts target thalamic sensorimotor connectivity

Following the failure of group-level analytical approaches, we sought to test whether effective contacts could be predicted at an individual level using biomarkers derived from diffusion MRI. We began by testing two tracts we hypothesised to be involved in mediating the effects of thalamic DBS: the cerebellothalamic tract (CTT, also known as the denticulo-rubro-thalamic tract), believed to be the putative target for tremor relief; and the pallido-thalamic tract (PTT), often regarded as one of the targets for relief of dystonic symptoms. Individual tractography revealed consistent tract reconstruction for both CTT (mean volume: 40,809mm^3^, mean length: 94.29mm, mean FA: 0.41) and PTT (mean volume: 37,662mm^3^, mean length: 53.0mm, mean FA: 0.44) (supplementary figure 1). However, effective contacts did not stimulate more of the CTT (effective: M 2.84 SD 2.98, ineffective M 2.14 SD 2.3, t(18)=0.58, *p*=.056) or PTT (effective: M 0.52 SD 0.82, ineffective M 0 SD 0, t(18)=, *p*=.01) than ineffective contacts (figure 4).

**Figure 4:**
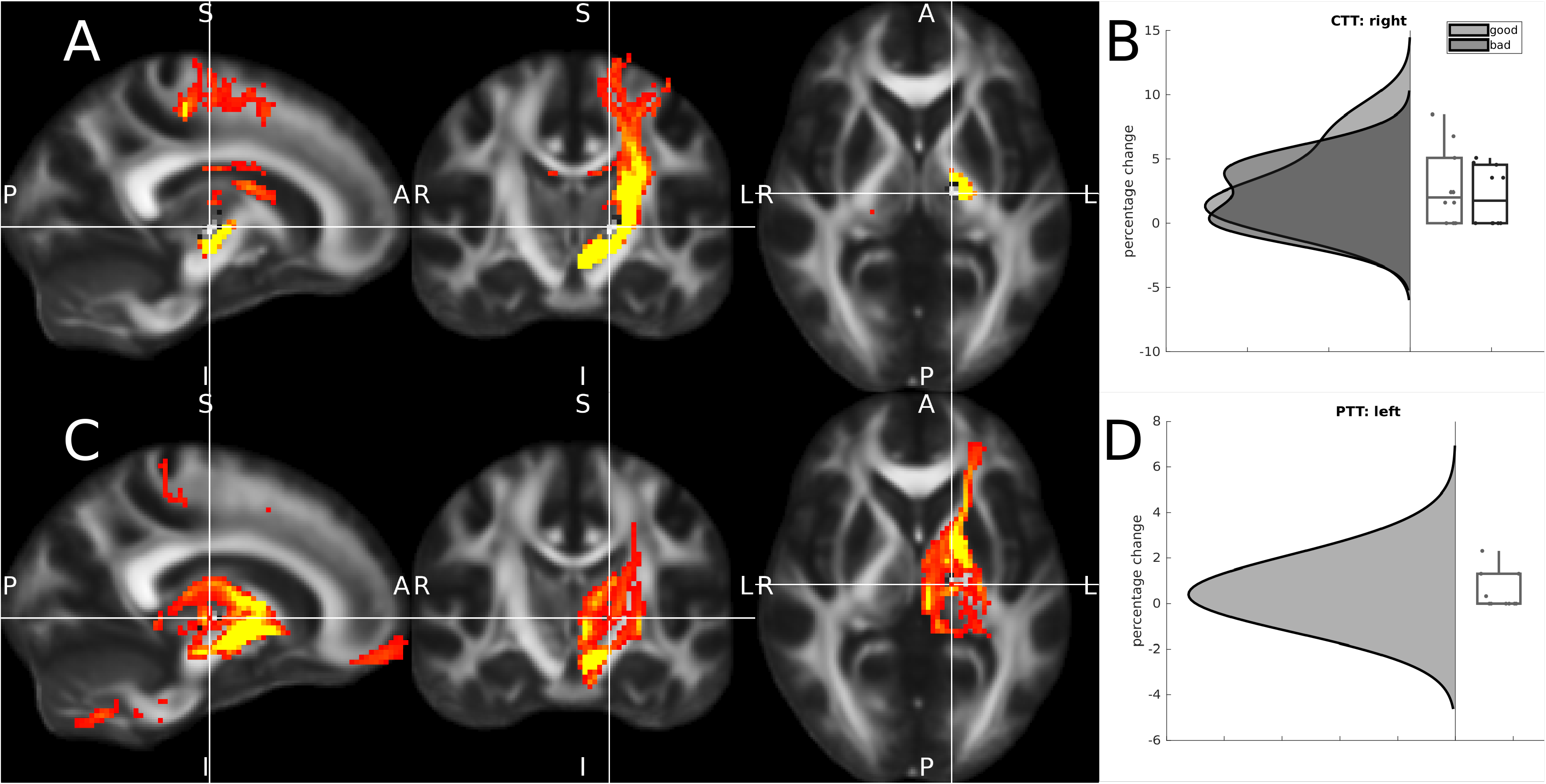
Tract-based Analyses. A: Group-level reconstruction of the right cerebello-thalamic tract (CTT) originating in the right dentate nucleus. B: Raincloud plots of effective versus ineffective contacts and stimulation of the CTT. C: Group-level reconstruction of the left pallido-thalamic tract (PTT). D: Raincloud plots of effective versus ineffective contacts and stimulation of the PTT.

Subsequently, we sought to understand if effective contacts involved the thalamus and its role in integrating sensorimotor activity and cortico-thalamic connectivity. Overall, consistency of thalamic segmentation methods was high (intra-class correlation co-efficient 0.74 to 0.75)(supplementary figure 2). For each segmentation approach, there were consistent biomarkers of effect (figure 5). For the clustering-based approach, effective contacts (M 2.5 SD 0.83) stimulated more of the thalamic segment connected to the primary motor cortex (M1) than ineffective contacts (M 0.79 SD 0.95, t(18)=4.29, *p*<0.001). For the hard-segmentation approach, effective contacts stimulated more of the thalamic segment connected to the sensorimotor target in the Yeo cortical atlas than ineffective contacts (effective: M 2.99 SD 0.96, ineffective: 1.78 SD 1.51, t(18)=2.13, *p*=.04). Furthermore, ineffective contacts stimulated more of the thalamic segment connected to the prefrontal region (effective: M 0 SD 0, ineffective M 7.37 SD 7.58, (18)=-3.08, *p*=.006). Finally, for k-means segmentation, this consistent pattern of segmentation stimulation was replicated, with effective contacts stimulating one segment (effective: M 1.89 SD 1.02, ineffective: M 0.52 SD 1.21, t(18)=2.75, *p*=0.013) and ineffective contacts stimulating another (effective: M 0 SD 0, ineffective: M 0.88 SD 0.68, t(18)=-4.09, p<0.001).

**Figure 5:**
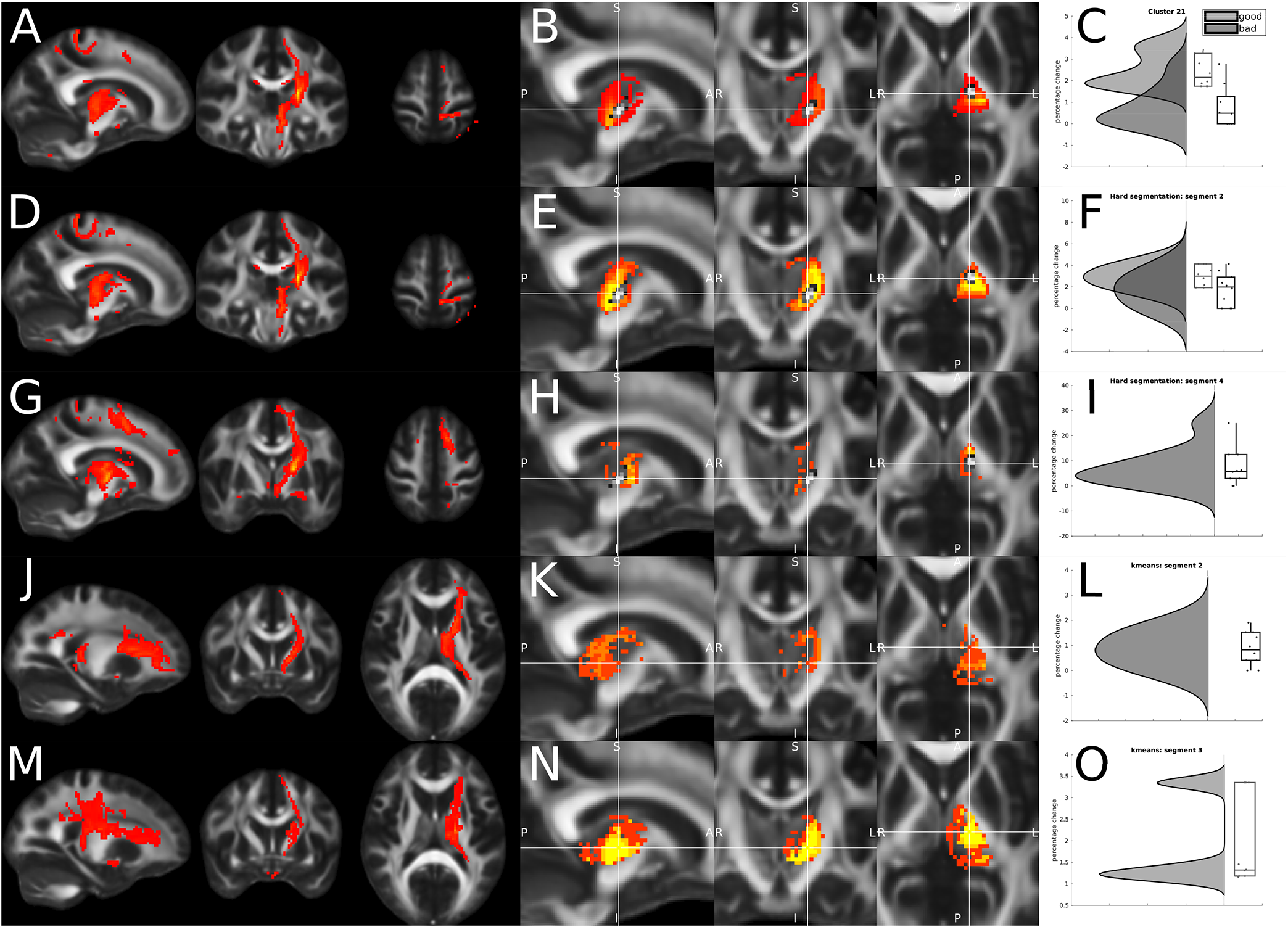
Segmentation Approach. A: Tractography from the thalamic segment most connected to the primary motor cortex (clustering-based approach to FreeSurfer cortical target 21). B: Corresponding individual thalamic segments from all participants. C: Raincloud plots demonstrating significantly more stimulation overlay for effective versus ineffective contacts. D: Tractography using the hard segmentation approach for connectivity to the sensory-motor cortex in the Yeo atlas. E: Corresponding individual thalamic segments from all participants. F: Raincloud plots demonstrating significantly more stimulation overlay for effective versus ineffective contacts. G: Tractography using the hard segmentation approach with connectivity to premotor areas in the Yeo atlas. H: Corresponding individual thalamic segments from all participants. I: Raincloud plots demonstrating stimulation overlay for ineffective but not effective contacts. J: Tractography from the thalamic segment derived by k-means segmentation demonstrating connectivity to prefrontal regions. K: Corresponding individual thalamic segments from all participants. L: Raincloud plots demonstrating stimulation overlay for ineffective but not effective contacts. M: Tractography from the thalamic segment derived by k-means segmentation demonstrating connectivity to sensory-motor regions. N: Corresponding individual thalamic segments from all participants. O: Raincloud plots demonstrating stimulation overlay only for effective contacts.

### IV. Final clinical stimulations are consistent with predicted targeting based on monopolar review but do not predict individual outcomes

Finally, we tested clinical outcomes at 12 months with the final clinical stimulations with each of our identified individual diffusion MRI biomarkers involved in thalamic segmentation. None of our biomarkers correlated with symptom improvement for V-RQOL and VHI (figure 6). However, final clinical stimulations did consistently include those thalamic areas identified already identified as effective based on monopolar review while avoiding thalamic areas identified as being ineffective (supplementary figure 3). In addition to the 5 main segments identified as being significantly stimulated on monopolar review, final clinical stimulations consistently activated 3 other targets including the right CTT, and segments connecting the primary sensory and secondary motor areas (using the clustering approach), suggesting that the final clinical effects involve a broader amalgamation of effects than on initial pure monopolar review.

**Figure 6:**
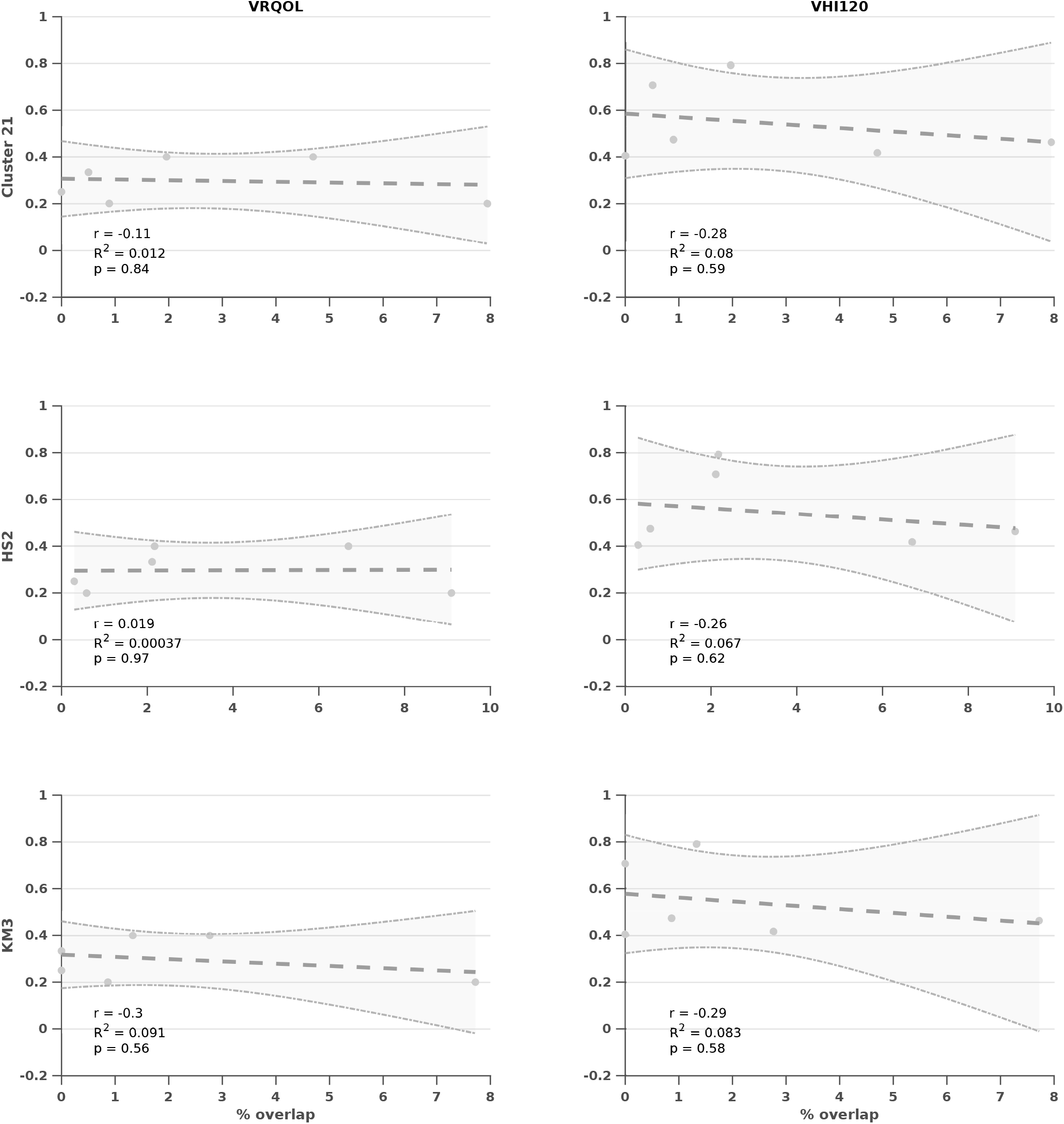
Clinical Outcomes and Stimulations. Clinical outcomes (dependent variables) for V-RQOL (first column) and VHI (second column) at 12 months with VATs using corresponding stimulation parameters at the time and the percentage stimulation (independent variables) of thalamic segments associated with effective contacts at monopolar review (figure 5) including: cluster-based segment to primary motor cortex (Cluster 21); hard-segmentation to sensory-motor cortex (HS2); and k-means segment to sensory-motor cortex (KM3). Plots show raw data, linear regression with confidence intervals, and corresponding statistical analyses. No significant findings were identified.

## DISCUSSION

We used diffusion MRI to identify pathways and individual biomarkers of response to thalamic DBS in SD. A consistent theme of individual biomarkers was a focus on thalamic sensorimotor connectivity independent of methodological or statistical approach. In contrast, group level targeting ‘sweet spots’ were not readily apparent. Finally, overall clinical outcomes involved stimulation of all the aforementioned targets identified by diffusion MRI but were not linearly correlated with clinical outcomes.

Neuroimaging methods are readily contributing to our understanding of the pathophysiological basis of spasmodic dysphonia. Reduced functional connectivity in the left sensorimotor cortex, inferior parietal cortex, putamen, right frontal operculum, and bilateral supplementary motor areas has been identified with resting-state functional MRI in people with spasmodic dysphonia compared with healthy controls(1,20) Dynamic causal modelling has elaborated on this hypothesis to suggest premotor-parietal-putaminal connectivity drives disorganised dystonic sensorimotor connectivity.(1) Somato-typical localisation of dopamine receptors within the striatum, identified through PET imaging, has identified alterations specific to the dystonic phenotype and provides a theory for linking symptomatology across dystonias.(2) These data are in keeping with our a priori hypothesis that DBS interacted with integration of sensorimotor connectivity within the thalamus. Our study further contributes to the hypothesis that the thalamus is a key contributor to the pathophysiology of spasmodic dysphonia, and that treatment efficacy is mediating through interactions with sensorimotor connectivity (albeit with the exact cellular mechanism still to be elucidated). Further studies testing the effects of pallidal and subthalamic DBS will be enlightening, although will likely rely on inference of their effects in other forms of dystonia given the paucity of studies specifically in spasmodic dysphonia.

Previous studies using diffusion MRI have identified numerous biomarkers for response of thalamic stimulation to tremor (its more traditional indication). These studies have used a range of MRI protocols, analysis methods and statistical techniques.(21) Our study was able to demonstrate consistent effects without complex sequences and independent of any single methodological approach, lending biological plausibility to the findings. We would encourage this approach of using a range of methods to test reproducibility and validity when applying neuroimaging methods in studies of DBS, which will ultimately facilitate identifying consistent effects and aid clinical translation.

A novel aspect of our study was utilising the effectiveness of monopolar review at very low voltages (0.5V) as our dependent variable. The rationale for this was that it increased the number of available data points, allowed precise localisation of the effective target within the thalamus, and was reliable in light of the objective measures of response. We would recommend this approach when participant numbers are constrained, particularly where targeting is highly accurate and leads to a significant overlap in the final stimulation volumes that can be untwined by investigating individual contacts. Consistency of monopolar review stimulation targets of both effective and ineffective thalamic segments with final clinical stimulations lends further clinical believability to this approach.

Future directions include using these novel diffusion MRI biomarkers to improve outcomes for people with spasmodic dysphonia treated with DBS. A safe and staged approach to individual targeting could be to leverage the technology of directional stimulation and pluri-contact electrodes to steer towards putative targets identified in the thalamus. If this is efficacious, the next step could be to consider individualised targeting to specific thalamic segments. Regarding neuroimaging methods, one particular avenue to explore is increased diffusion directions to allow more tractography to orofacial regions of the sensory-motor cortex which have a higher angular resolution as they diverge in the centrum semi-ovale. Finally, this work will inherently rely on large-scale co-operative research, given the relative paucity in numbers of people treated, in order to validate the reproducibility of this approach.

The main limitation of this study is the relatively low number of participants involved. However, this is still the largest series of people with spasmodic dysphonia having DBS, and we have adapted to this challenge by using a novel approach utilising the results of the monopolar review. Neuroimaging methods, specifically sequence protocols and analytical techniques, could be increased in their complexity. However, a strength of this study is the application of methods that are more readily translatable to routine clinical practice. Finally, an area not directly touched upon in this study is the aspect of optimal patient selection and objective diagnostic criteria. By the time DBS is considered people with spasmodic dysphonia have often had many years of symptoms and therapy, adding to diagnostic and prognostic complexity. Therefore, selection of people most likely to benefit should be seen as complimentary to this work seeking to maximise the benefit of those who do have DBS.

## CONCLUSIONS

Thalamic DBS improves symptoms of spasmodic dysphonia both at 12 months and at initial monopolar review. A convergence of diffusion MRI analyses highlight a critical role of sensorimotor connectivity within the thalamus as a key site of treatment efficacy and can serve as novel biomarkers for targeting. Future work could leverage directional DBS electrodes to steer current towards these diffusion MRI biomarkers and test the hypotheses stimulation of sensorimotor connectivity within the thalamus drives symptom improvement.

## Data Availability

All data produced in the present study are available upon reasonable request to the authors.

## ACKNOWLEDGEMENTS

Nil

## Abbreviations

AC: anterior commissure
ANT: advanced normalization tools
FA: fractional anisotropy
FDR: false discovery rate
FGATIR: Fast Gray Matter Acquisition T1 Inversion Recovery
Hz: Hertz
M: mean
MCP: mid-commissural point
ms: millisecond
PaCER: Precise and Accurate Electrode Reconstruction
PC: posterior commissure
SD: standard deviation
V: Volts, VAT volume of activated tissue
VLdVLv: Ventral Lateral dorsal / Ventral Lateral ventral
VIM: ventral intermediate nucleus

## FIGURES

**SF1: Individual Tractography**

Raw individual tractography data for the cerebello-thalamic tract (CTT) in the first column and the pallido-thalamic tract (PTT) in the second column for individual participants (rows).

**SF2: Individual Segmentation**

Raw individual segmentation data for the clustering-based approach for FreeSurfer derived cortical targets (first column), hard-segmentation approach to cortical targets in the Yeo atlas (second column), and data-driven k-means approach to an individual cortical grey-matter mask (third column). Each row is data for an individual participant. Intra-class correlation coefficients ranged from 0.74 to 0.75.

**SF3: Overall Stimulation**

Raincloud plots of clinical VATs at 12 months and percentage stimulation of tractography (top row), clustering-based segmentation analysis (second row), hard-segmentation analysis (third row), and k-means segmentation (bottom row). Only segments consistently stimulated are shown which include all those identified on monopolar review as well as spread into additional right CTT and clustering-based segments to the primary sensory and secondary motor areas.

