## Supplementary figures and images for "Deep brain stimulation improves symptoms of spasmodic dysphonia through targeting of thalamic sensorimotor connectivity"

### Supplementary Figure 1

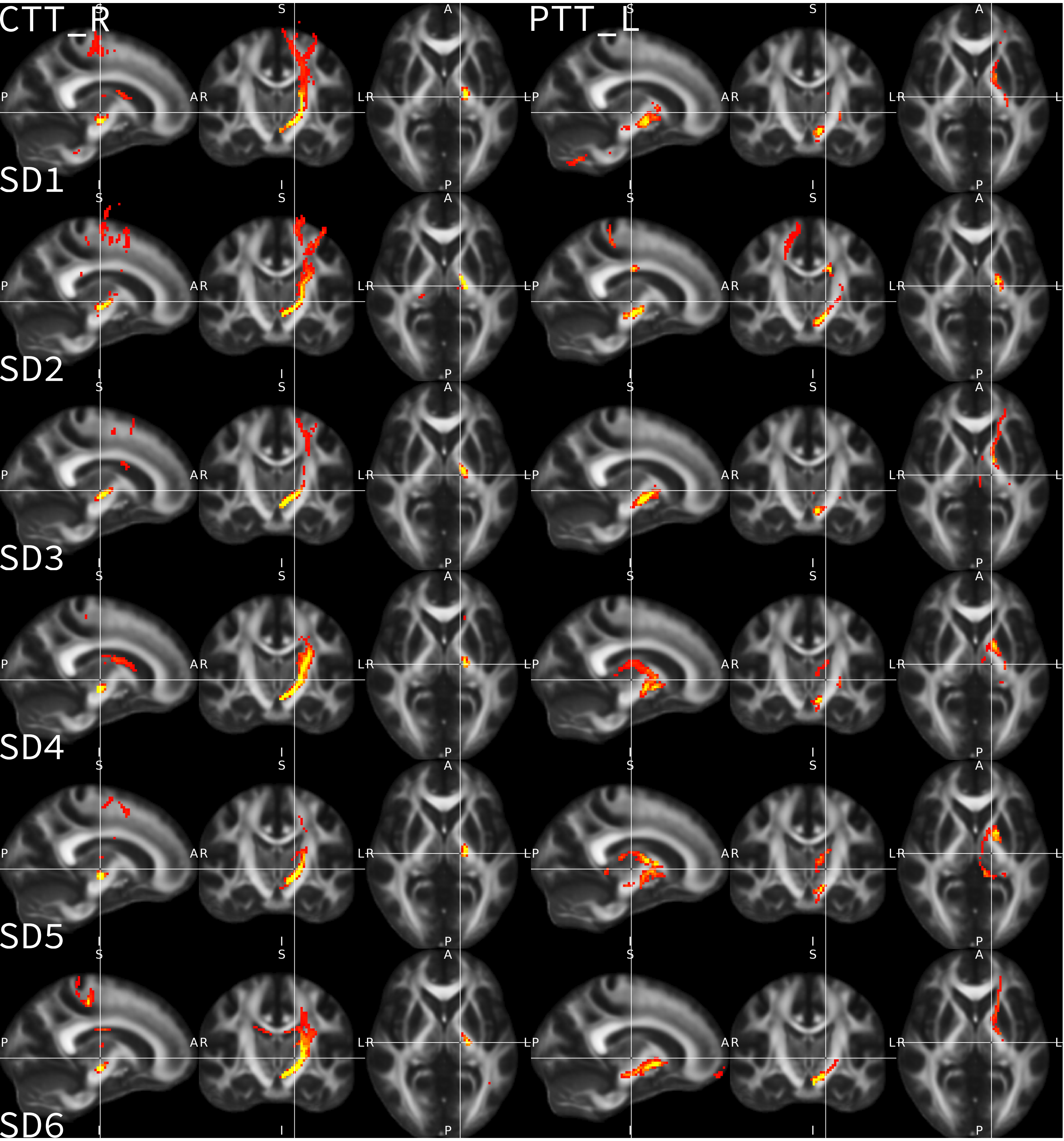

### Supplementary Figure 2

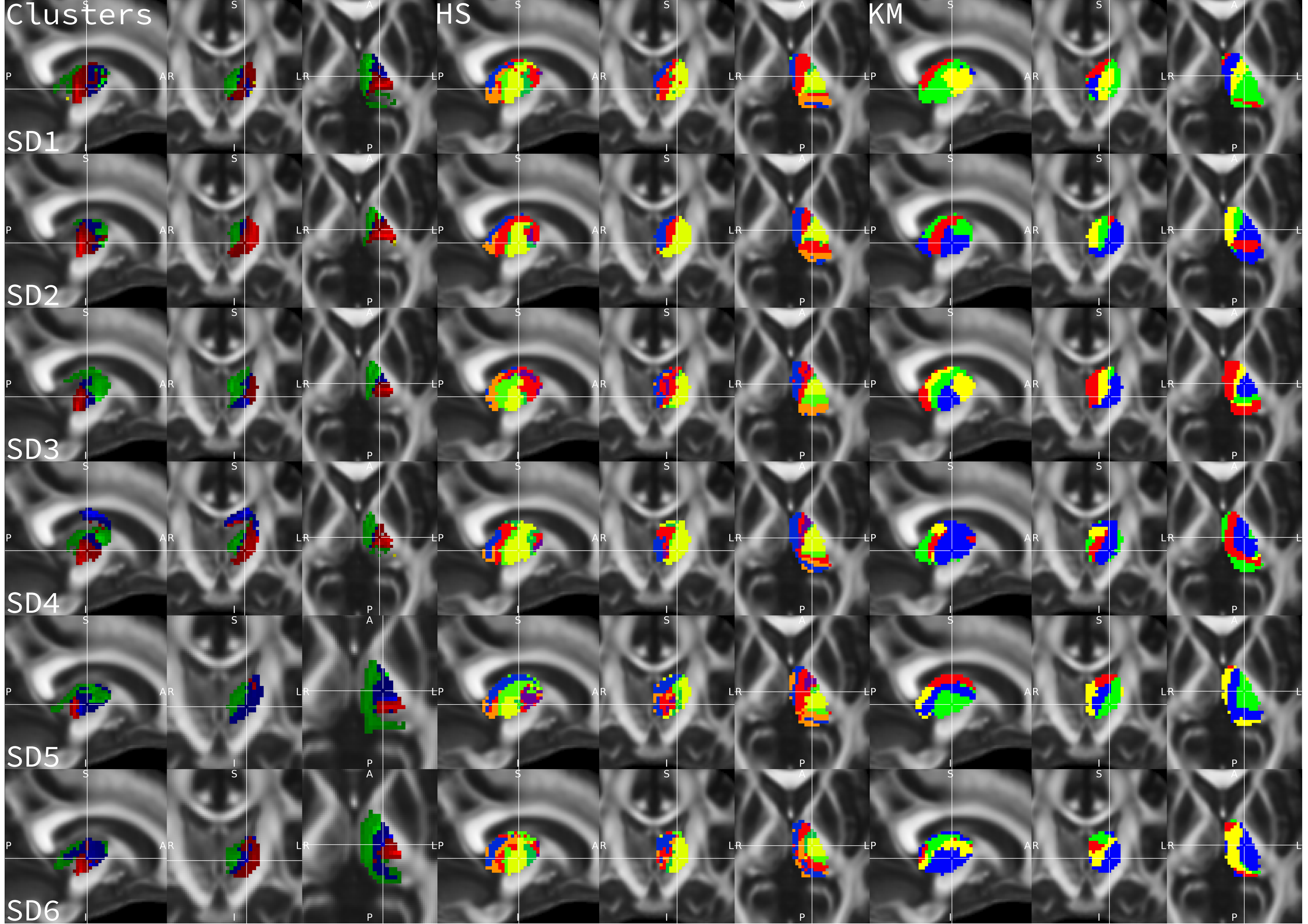

### Supplementary Figure 3

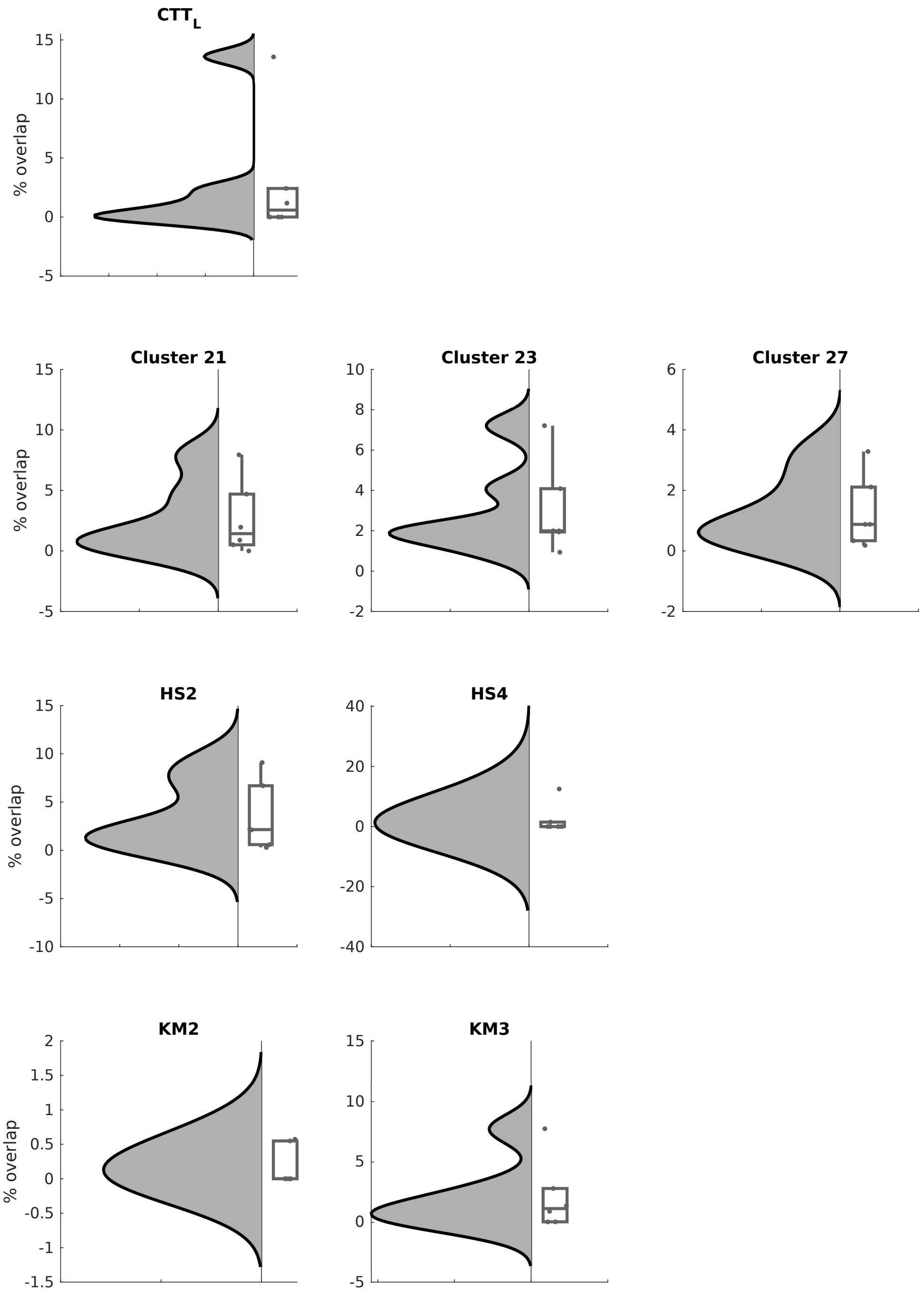
